# Availability and Utilization of Sexual and Reproductive Health Services among Adolescents of Godawari Municipality, Nepal: a Cross sectional Study

**DOI:** 10.1101/2024.05.08.24307068

**Authors:** Laxmi Gautam, Aastha Maharjan, Harikrishna Bhattarai, Sujan Gautam

## Abstract

**Background:** Adolescent sexual and reproductive health is getting due attention in developing countries because investing among adolescent can bring multiple benefits. This study aims to assess the availability, utilization and explore different factors affecting the utilization of SRH services by adolescents of Godawari Municipality.

**Methods:** cross-sectional, mixed-method study was conducted using the Anderson’s model of health service utilization in 3 government secondary schools and three local health institutions in Godawari Municipality. A self-administered questionnaire was used among adolescents aged 15- 19 (n = 416) and Key Informants Interview (KII) and observation were done in health institutions. Quantitative data was analyzed in Statistical Package for Social Sciences (SPSS) version 16 and thematic analysis was done for qualitative data.

**Results:** The mean age of the respondents was 17.31±0.99 years. Nearly one-third of them (30.63%) had not heard about Adolescents Friendly Health Services. Knowledge of ASRH, AFHS (AOR=2.814, 95% C.I=1.129-7.013) and conversation with parents (AOR=2.069, 95% C.I=1.094-3.912), availability of ASRH (AOR=2.801, 95% C.I=0.810-9.687) had significant relationship with utilization of Health Services. The adolescents’ perceived feeling of the need for ASRH services is significantly associated with the utilization of ASRH services (p=< .0001). Only 13.22% of them had ever used ASRH services and the reasons for not using services was the lack of realization of the need of services (60.39%) while 14.55% of them felt that privacy was not maintained at all. The KII found the number of adolescents visiting Health Facility (HF) was very less and those visiting for the SRH related services were rare. The observation showed none of the HF met the adolescents friendly criteria set by government even when they were once certified.

**Conclusion:** Knowledge of respondents and their parents as well as availability of the services had significant role in the utilization of ASRH service by the adolescents. Awareness about the importance of health care services among the adolescent and the availability of services is essential.

## Introduction

The World Health Organization (WHO) definition officially designates an adolescent as someone between the ages of 10 and 19. Adolescents experience rapid physical, cognitive and psychosocial growth. This affects how they feel, think, make decisions, and interact with the world around them.(1) ASRH refers to the physical and emotional wellbeing of adolescents and includes their ability to remain free from unwanted pregnancy, unsafe abortion, Sexually Transmitted Infections (including HIV/AIDS), and all forms of sexual violence and coercion.(2) The health needs of young people are special so they need special services called Adolescents Friendly Health Services (AFHS), which are defined by the friendly environment and favorable condition where adolescents get access to ASRH services in accessible, equitable, acceptable, appropriate, and effective manner.(3) However, little information exists regarding in what Nepalese adolescents perceive as their SRH needs, their service utilization patterns and factors which act as barriers or facilitates use of existing services.(4)

Adolescents in Nepal do not have adequate access to information and access to services which is further exacerbated by the little sex education in schools and hardly open discussion about sex and sexuality in families and society.(5) There are limited sessions on adolescent’s sexual and reproductive health in school curriculum. Therefore, adolescents are unable to receive complete information about their physiological, psychological and emotional changes that occur during puberty.(6) To meet the health needs of adolescents, the Nepal government launched a national program in 2010 to provide adolescent-friendly sexual and reproductive health services as part of its five-year health sector plans. (7) Nepal Demographic Health Survey (NDHS) 2022 prevailed that 14% of women age 15–19 have ever been pregnant, 10% have had a live birth, and 2% have had a pregnancy loss. Two percent each of women and men age 15–19 had sexual intercourse before age 15. Three percent of women age 15–19 were married by age 15, and 1% had been pregnant by that age. (8) Although SRH education and services are provided to young people, unplanned pregnancy, unsafe abortion, sexually transmitted infections are commonly existed among adolescents in Nepal, indicating underutilization of sexual health services by young people. Health facilities have apparently failed to provide young people with specialized sexual health education and services.(9) However, despite the interest of young people in obtaining relevant information and friendly services, the provision of sexual and reproductive health services in Nepal is very inadequate. However, little information exists at the national level regarding what Nepalese adolescents perceive as their SRH needs, their service utilization patterns and factors which act as barriers or facilitates use of existing services.(10) Global evidence shows that programs such as comprehensive sexuality education and youth-friendly services have a positive influence on SRH service utilization.(11) Poor sexual and reproductive health knowledge, the lack of youth-friendly services, lack of confidentiality of services, experiences of shame, and health care provider attitudes also have an influence on SRH service utilization by adolescents.(12) Correct evidence is crucial to properly plan SRH services for Nepalese adolescents. Amongst the many means to measure adolescents’ needs, their own reports of health behaviors and lifestyles and utilization of health services are especially valuable. So, this study aims to assess the availability and utilization of ASRH services and explores different factors affecting the utilization of the services related to individual, health care institution and service providers.

## Materials and Methods

A descriptive cross-sectional study was conducted to assess the availability and utilization of ASRH services by using quantitative and qualitative methods from November 2021 to January 2022. The dependent variable was the availability and utilization of SRH services by adolescents and the independent variables were socio-demographic factors, health facility, service provider-related factors, SRH information-related factors, and so on.

For the quantitative study, 416 school-going adolescents aged 15 to 19, studying in classes 11 and 12 of three government secondary schools were selected in Godawari Municipality, Lalitpur. The study was conducted in three government schools selected randomly from 10 schools which were located in different 3 wards wards out of 14 wards of Godawari Municipality. The sample size was calculated using the formula for infinite population developed by Cochran with a 95% CI, 5% allowable error, and a non-response rate of 10%. The schools were selected by lottery method and the number of students of each school and class was selected by Probability Proportionate Sampling (PPS). The individual participants were selected by lottery method. For the qualitative study three local health institutions and the health service provider of those institutions were selected purposively that were located around the selected schools among four adolescent-friendly certified health institutions available in the municipality.

During data collection, permission was taken from the Municipality health section and education section, and information about schools and health institutions was gathered. The information about the adolescents-friendly certified health institutions of the municipality was collected from the family welfare division. Then, schools were contacted and permission was taken from the school administration before collecting data from the students.

Quantitative data was collected using a self-administered, semi-structured questionnaire and the qualitative data by a face-to-face interview using an interview guide. The health institutions were observed using a checklist. The data collection tools were developed after an extensive literature review, based on various previously conducted studies and were pretested in Kageswori Manohara Municipality. The observation checklist was adapted from the Government of Nepal’s criteria for Adolescent Friendly Health Institutions. Pretesting was done among 10% of the sample size and in one health institution to check instruments and a few modifications were made. The questionnaire was translated into the Nepali language.

For the analysis of quantitative data, IBM-SPSS software version 16 was used. Descriptive statistics were used to study the characteristics of variables and Chi-square test was used to test the association and logistic regression modeling was done. The information from the interviews was noted and recorded with the permission of the interviewee. They were transcribed into Nepali and then translated into English, which was analyzed through the process of thematic analysis by understanding the main themes of information and coding them into similar themes categories. The important information from the observation checklist was noted.

### Ethical Statement

The proposal was approved by the Institutional Review Committee (IRC), at the Manmohan Memorial Institute of Health Science (MMIHS-IRC 619). Study was conducted from 26^th^ November 2021 to 17^th^ January 2022.Written Informed consent was taken with each respondent before data collection for quantitative study while verbal informed consent was taken for qualitative study. In case of participants below the age of 18 permission was taken from school principle and their respective class teachers and then assent was taken from each of them. Participation was voluntary and all ethical considerations, including confidentiality and privacy were maintained throughout the study process.

## Result

The mean age of the respondents was 17.31±0.99 years. More than half of the participants were 15-17 years old (57.87%), female (55.05%), and were studying in class 12 (55.67%). Only 13.83% of the participants were married or in a relationship and the relationship status was statistically associated (p=0.024) with utilization of ASRH. Only about one-third of the respondents (30.53%) had conversation with the parents regarding different matters of ASRH. However, factors such as age, sex, the class they study in were not significantly associated with utilization. Regarding the educational status of parents, 13.89% of them had illiterate father which was 28.15 % in case of mother.

Almost all (99.28%) of the respondents knew about or were aware of ASRH. Nearly one-third of them (30.63%) had not heard about Adolescents Friendly Health Services. More than three-quarters (77.40%) of them knew about any health institutions that provided ASRH services in their area. Knowledge of ASRH, AFHS and conversation with parents had significant relationship with utilization of AHS utilization. (Table 1)

**Table 1:** Predisposing and enabling factors of ASRHS utilization.

| Variables | Utilization of ASRHS |  | Total |  | P value |
| --- | --- | --- | --- | --- | --- |
|  | Yes (%) | No (%) | Frequency | % |  |
| **Predisposing factors** |  |  |  |  |  |
| Age: Mean ±SD = 17.31 ± 0.99 years (n=413) | | | | | |
| $\leq 17$ years | 30 (12.60) | 209 (87.40) | 239 | 57.87 | 0.592 |
| $> 17$ years | 25 (14.40) | 149 (85.60) | 174 | 42.13 | |
| **Sex** |  |  |  |  |  |
| Male | 26 (13.90) | 161 (86.10) | 187 | 44.95 | 0.710 |
| Female | 29 (12.70) | 200 (87.30) | 229 | 55.05 |  |
| **Level of Education** |  |  |  |  |  |
| Grade 11 | 24 (12.70) | 165 (87.30) | 189 | 45.43 | 0.774 |
| Grade 12 | 31 (13.70) | 196 (86.30) | 227 | 55.67 |  |
| **Religion** |  |  |  |  |  |
| Hindu | 39 (12.80) | 266 (87.20) | 305 | 73.32 | 0.665 |
| Others | 16 (14.40) | 95 (85.60) | 111 | 26.68 |  |
| **Relationship status (n=412)** |  |  |  |  |  |
| Married/ In Relationship | 13 (22.80) | 44 (77.20) | 57 | 13.83 | 0.024 |
| Single/ Others | 42 (11.80) | 313 (88.20) | 355 | 86.27 |  |
| **Enabling Factors** |  |  |  |  |  |
| **Father's Education (n=403)** |  |  |  |  |  |
| Illiterate | 8 (14.30) | 48 (85.70) | 56 | 13.89 |  |
| Primary school | 12 (10.30) | 105 (89.70) | 117 | 29.03 | 0.756 |
| Secondary school | 22 (15.30) | 122 (84.70) | 144 | 35.73 |  |
| High school | 6 (16.70) | 30 (83.30) | 36 | 8.93 |  |
| Literate only | 6 (12.00) | 44 (88.00) | 50 | 12.40 |  |
| **Mother's Education (n=414)** |  |  |  |  |  |
| Illiterate | 15 (87.10) | 101 (12.90) | 116 | 28.15 | 0.900 |
| Primary school | 14 (14.10) | 85 (85.90) | 99 | 23.91 |  |
| Secondary school | 10 (12.20) | 72 (87.80) | 82 | 1.98 |  |
| High school | 1 (6.30) | 15 (93.80) | 16 | 3.86 |  |
| Literate only | 15 (14.90) | 86 (85.10) | 101 | 24.40 |  |
| **Heard About ASRH** |  |  |  |  |  |
| Yes | 55 (13.30) | 358 (87.00) | 413 | 99.28 | 1.000* |
| No | 0 (0.00) | 3 (100.00) | 3 | 0.72 |  |
| **Known the Institutions providing ASRH** |  |  |  |  |  |
| Yes | 52 (16.10) | 270 (83.90) | 322 | 77.40 | 0.004* |
| No | 0 (0.00) | 25 (100.00) | 25 | 6.00 |  |
| Don't Know | 3 (4.30) | 66 (95.70) | 69 | 16.60 |  |
| **Heard about AFHS** |  |  |  |  |  |
| Yes | 49 (16.90) | 240 (83.10) | 289 | 69.47 | 0.001 |
| No | 6 (4.70) | 121 (95.30) | 127 | 30.63 |  |

| **Conversation with parents about ASRH** |  |  |  |  |  |
| --- | --- | --- | --- | --- | --- |
| Yes | 26 (20.50) | 101 (79.50) | 129 | 30.53 | 0.004 |
| No | 29 (10.00) | 260 (90.00) | 285 | 69.47 |  |
| **Parents support in use of services (n=413)** |  |  |  |  |  |
| Yes | 46 (14.60) | 270 (85.40) | 316 | 76.51 | 0.181 |
| No | 9 (9.30) | 88 (90.70) | 97 | 23.49 |  |

Almost half of the respondents reported that they had felt the need for ASRH services (48.56%) until now. The adolescents’ perceived feeling of the need for ASRH services is significantly associated with the utilization of ASRH services (p=< 0.0001). Majority of the respondents (88.43%) felt the need for separate and specific services for adolescents. Nearly two third (65.62%) thought that their decision on ASRH service utilization would be affected by the gender of the service provider. However, their perception of the need for separate and specific services for adolescents and the effect of the sex of the service provider were not significantly associated with the utilization of ASRH services. Only two third (66.59%) of the respondents thought they would use the service in the future if needed. Those who were willing to use ASRH service if needed in the future were significantly associated with the utilization of services. The respondents who intended to use the services in future were 5.882 times more likely to utilize ASRH services compared to those who were not sure of it (COR= 5.882, 95% C.I= 2.070-16.717).

For the more than 2/3^rd^ of respondents (68.94%), the nearest health institution was less than 30 minutes walking distance from their home. However, information/ awareness about ASRH and the distance to the health institution from home were not significantly associated with the utilization. Majority of the respondents (82.13%) reported the need of convenient opening hour which was significantly associated with the utilization of ASRHS. (Table 2)

**Table 2:** Need and External Environmental factors of ASRHS utilization.

| Variables | Utilization of SRHS |  | Total |  | P value |
| --- | --- | --- | --- | --- | --- |
|  | Yes (%) | No (%) |  |  |  |
| **Need Factors** |  |  |  |  |  |
| **Felt need of service** |  |  |  |  |  |
| Yes | 55 (100.00) | 147(40.70) | 202 | 48.56 | < .0001 |
| No | 0 (0.00) | 214 (59.30) | 214 | 51.44 |  |
| **Perceived need of separate service (n=415)** |  |  |  |  |  |
| Yes | 48 (13.10) | 319 (86.90) | 367 | 88.43 | 0.773 |
| No | 7 (14.60) | 41 (85.40) | 48 | 1157 |  |
| **Use in future if needed** |  |  |  |  |  |
| Yes | 49 (17.80) | 227 (82.20) | 276 | 66.35 | 0.001 |
| No | 2 (7.40) | 25 (92.60) | 27 | 6.49 |  |
| Don't Know | 4 (3.50) | 109 (96.50) | 113 | 27.16 |  |
| **External environmental factors** |  |  |  |  |  |
| **Distance to health institution (n=322)** |  |  |  |  |  |
| <30 minutes walking distance | 34 (15.30) | 188 (84.70) | 222 | 68.94 | 0.641 |
| 30 minutes to 1 hour walking distance | 11 (16.20) | 57 (83.80) | 68 | 21.12 |  |
| >1 hour walking distance | 7 (21.90) | 25 (78.10) | 32 | 9.94 |  |
| **Necessity of Convenient Opening hours of institution** |  |  |  |  |  |
| Yes | 53 (15.60) | 287 (84.40) | 340 | 82.13 | 0.010 |
| No | 2 (2.70) | 72 (97.30) | 74 | 17.87 |  |
| **Perceived Community support** |  |  |  |  |  |
| Yes | 38 (15.70) | 204 (84.30) | 242 | 58.31 | 0.082 |
| No | 17 (9.80) | 156 (90.20) | 173 | 41.69 |  |
| **Perceived Religious and cultural beliefs/Barriers** |  |  |  |  |  |
| Yes | 20 (14.40) | 119 (85.60) | 139 | 33.41 | 0.619 |
| No | 35 (12.60) | 242 (87.40) | 277 | 66.59 |  |
| **Perceived Effect of sex of service provider (n=413)** |  |  |  |  |  |
| Yes | 36 (13.30) | 235 (86.70) | 271 | 65.62 | 0.862 |
| No | 18 (12.70) | 124 (87.30) | 142 | 34.38 |  |

The respondents who had conversation with their parents about ASRH were 2.308 times more likely to utilize ASRH services than the respondents who did not have conversation with parents on ASRH related matters (COR=2.308, 95% C.I= 1.296, 4.110). The respondents who knew about AFHS were 4.117 times more likely to utilize services than the respondents who were not aware about AFHS (COR=4.117, 95% C.I= 1.716, 9.881). The respondents who knew about the health institutions providing ASRH services are 4.237 more likely to use services compared to the ones who did not know about it (COR=4.237, 95% C.I= 1.283 – 13.990). Because of the no respondents who use ASRHS without the felt need of services and Knowledge about Institutions providing ASRH, logistic regression was not applicable even though they were significant factors in utilizing ASRHSs. (Table 3)

**Table 3:** Factors associated with utilization of ASRH services by using multiple logistic regression.

| Factor | Utilization of services |  | 95%CI<br>(lower-upper<br>limit) | AOR | p-<br>value |
| --- | --- | --- | --- | --- | --- |
|  | Yes<br>n (%) | No<br>n (%) |  |  |  |
| Conversation with parents about ASRH |  |  |  |  |  |
| Yes | 26 (20.50) | 101 (79.50) | 1.094 – 3.912 | 2.069 | 0.25 |
| No | 29 (10.00) | 260 (90.00) | Ref |  |  |
| Heard about AFHS |  |  |  |  |  |
| Yes | 49 (16.90) | 240 (83.10) | 1.129 – 7.013 | 2.814 | 0.026 |
| No | 6 (4.70) | 121 (95.30) | Ref |  |  |

| **Availability of health institution providing ASRH services** |  |  |  |  |  |
| --- | --- | --- | --- | --- | --- |
| Yes | 52 (16.10) | 270 (83.90) | 0.810 – 9.687 | 2.801 | 0.104 |
| No | 0 (0.00) | 25 (100.00) |  | 0.00 | 0.998 |
| Don't Know | 3 (4.30) | 66 (95.70) | Ref |  |  |
| **Comfortable opening hours** |  |  |  |  |  |
| Yes | 53 (15.60) | 287 (84.40) | Ref |  |  |
| No | 2 (2.70) | 72 (97.30) | 0.039 – 0.716 | 0.167 | 0.16 |
| **Willingness to Use service in future** |  |  |  |  |  |
| Yes | 49 (17.80) | 227 (82.20) | 1.623 – 13.997 | 4.766 | 0.005 |
| No | 2 (7.40) | 25 (92.60) | 0.323 - 12.278 | 1.993 | 0.457 |
| Don't know | 4 (3.50) | 109 (96.50) | Ref |  |  |

### Characteristics related to Utilization of services by Respondents

The utilization of the ASRH by the respondents. Very few of them (13.22%) had ever used ASRH services till now among them maximum used those services in government health institutions (69.01%). Most of them got general health checkup and general health information counseling services and condom collection and family planning services. The reasons for not using services were, mostly because they did not think they needed the service (60.39%). Other reasons were them not being interested in using such services (38.23%), ashamed to ask for services (26.31%) and not knowing where to get such services (20.22%). Some of them (7.48%) also responded that opposite sex of the service providers was also the reason. Around half of them (52.73%) had paid for the services. Most of them (81.82%) had seen IEC materials in institute. Around 40.00% did not have to wait or only waited <30 minutes for services. Three fourth (70.91%) of them were comfortable to ask the questions. Almost all (90.91%) said that they were given proper attention by health workers. Less than one fourth (14.55%) of them felt that privacy was not maintained at all. Almost all of them (94.55%) would recommend others to go to the health institutions. (Table 4)

**Table 4:**
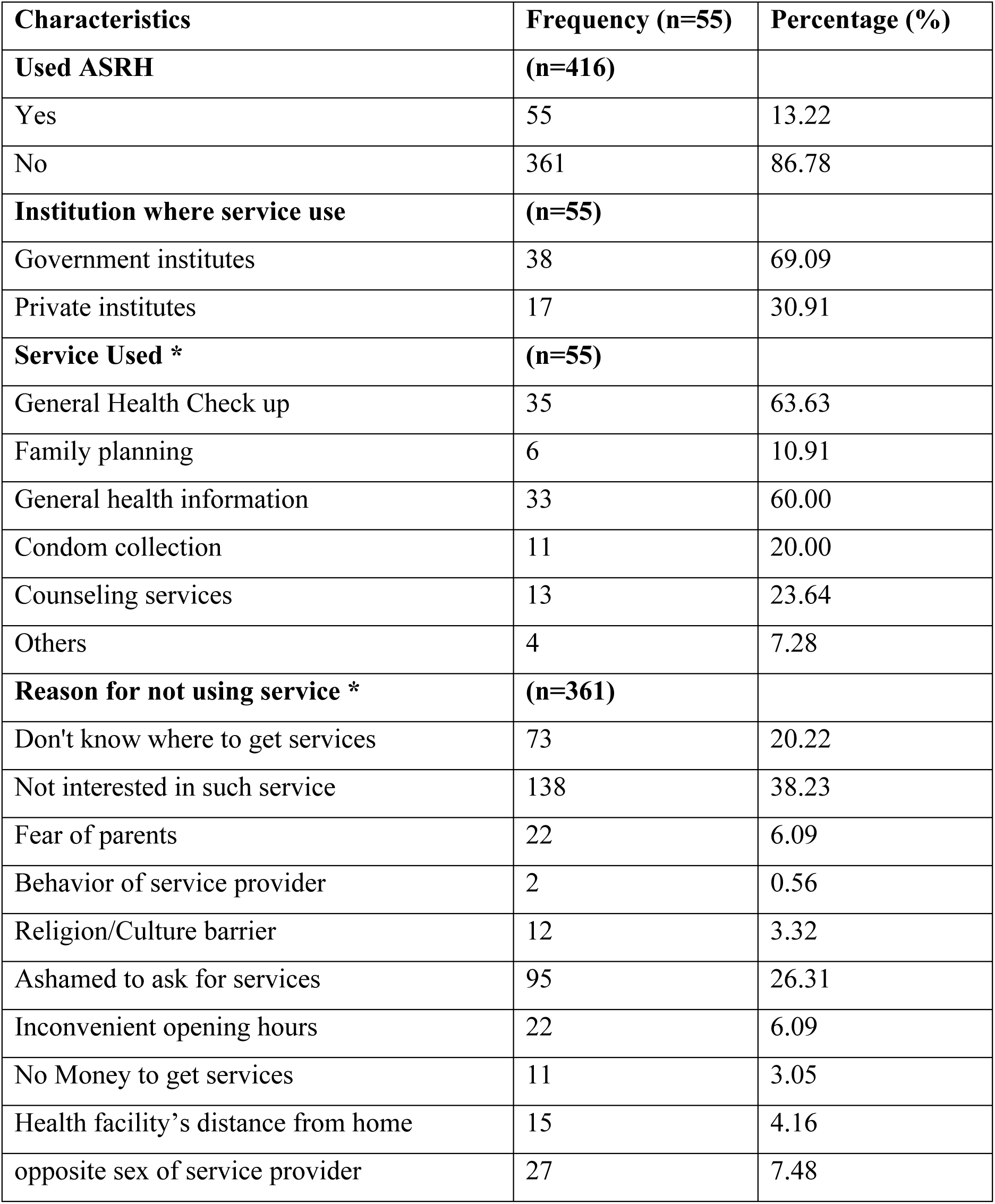
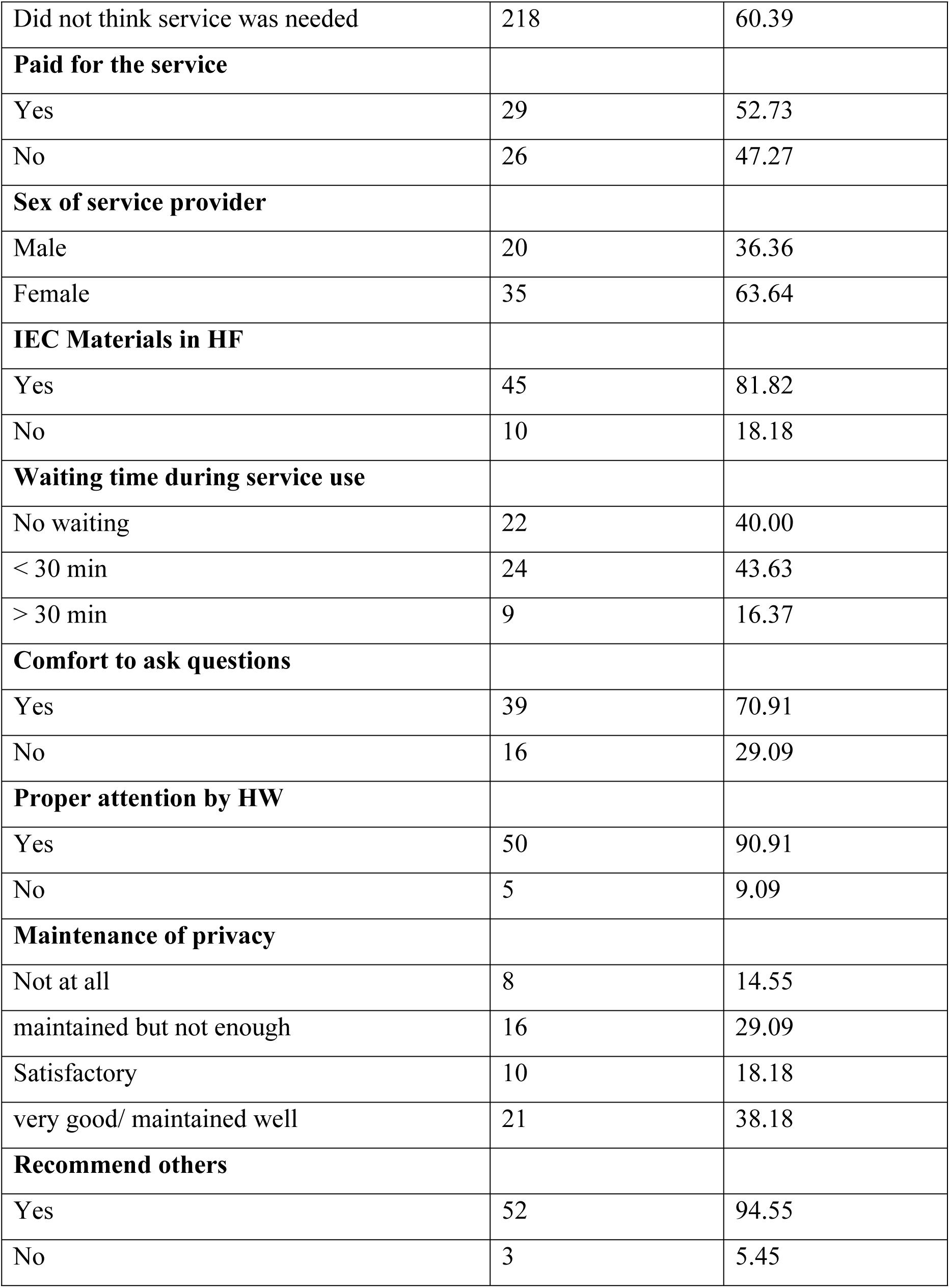
Characteristics related to experience during Utilization of ASRH services.

| **Characteristics** | **Frequency (n=55)** | **Percentage (%)** |
| --- | --- | --- |
| **Used ASRH** | **(n=416)** |  |
| Yes | 55 | 13.22 |
| No | 361 | 86.78 |
| **Institution where service use** | **(n=55)** |  |
| Government institutes | 38 | 69.09 |
| Private institutes | 17 | 30.91 |
| **Service Used \*** | **(n=55)** |  |
| General Health Check up | 35 | 63.63 |
| Family planning | 6 | 10.91 |
| General health information | 33 | 60.00 |
| Condom collection | 11 | 20.00 |
| Counseling services | 13 | 23.64 |
| Others | 4 | 7.28 |
| **Reason for not using service \*** | **(n=361)** |  |
| Don't know where to get services | 73 | 20.22 |
| Not interested in such service | 138 | 38.23 |
| Fear of parents | 22 | 6.09 |
| Behavior of service provider | 2 | 0.56 |
| Religion/Culture barrier | 12 | 3.32 |
| Ashamed to ask for services | 95 | 26.31 |
| Inconvenient opening hours | 22 | 6.09 |
| No Money to get services | 11 | 3.05 |
| Health facility's distance from home | 15 | 4.16 |
| opposite sex of service provider | 27 | 7.48 |
| Did not think service was needed | 218 | 60.39 |
| **Paid for the service** |  |  |
| Yes | 29 | 52.73 |
| No | 26 | 47.27 |
| **Sex of service provider** |  |  |
| Male | 20 | 36.36 |
| Female | 35 | 63.64 |
| **IEC Materials in HF** |  |  |
| Yes | 45 | 81.82 |
| No | 10 | 18.18 |
| **Waiting time during service use** |  |  |
| No waiting | 22 | 40.00 |
| < 30 min | 24 | 43.63 |
| > 30 min | 9 | 16.37 |
| **Comfort to ask questions** |  |  |
| Yes | 39 | 70.91 |
| No | 16 | 29.09 |
| **Proper attention by HW** |  |  |
| Yes | 50 | 90.91 |
| No | 5 | 9.09 |
| **Maintenance of privacy** |  |  |
| Not at all | 8 | 14.55 |
| maintained but not enough | 16 | 29.09 |
| Satisfactory | 10 | 18.18 |
| very good/ maintained well | 21 | 38.18 |
| **Recommend others** |  |  |
| Yes | 52 | 94.55 |
| No | 3 | 5.45 |

The KII among health workers mentioned that the number of adolescents visiting health institutions was less and those visiting for SRH-related services were rare. However, there was no separate recording system for adolescents, so accurate information about the number of adolescent visits was unavailable. The ones visiting the institutions would come for general reasons like fever, first-aid, or other OPD services. The barriers identified in the delivery of services to adolescents were that the infrastructure was not adolescent-friendly such as separate rooms, proper waiting spaces, the unavailability of appropriate IEC materials, and the unavailability of trained health workers to provide specific ASRH services. The health institutions only provided limited services like General health check-ups, health information, counseling services, family planning services, and pregnancy tests. They were not able to give comprehensive services to adolescents.

While observing the health institutions, all three were once certified as adolescent-friendly health institutions, but currently, they did not meet any specific criteria of being adolescents friendly. There was no supply of ASRH-specific IEC materials, no trained health workers, no adolescent job aid was available, none of them had displayed the AFS logo, reporting in AFs format was null, and involvement of the adolescents in HFOMC was not in practice. Criteria met to some extent were the convenient location of the health institution, toilets with clean water and dustbin, maintenance of privacy as far as possible while providing service, and counseling. Even though those health institutions were once certified as AF institutions, due to a lack of proper monitoring and follow-up, they could not maintain that status anymore. (Table 5)

**Table 5:**
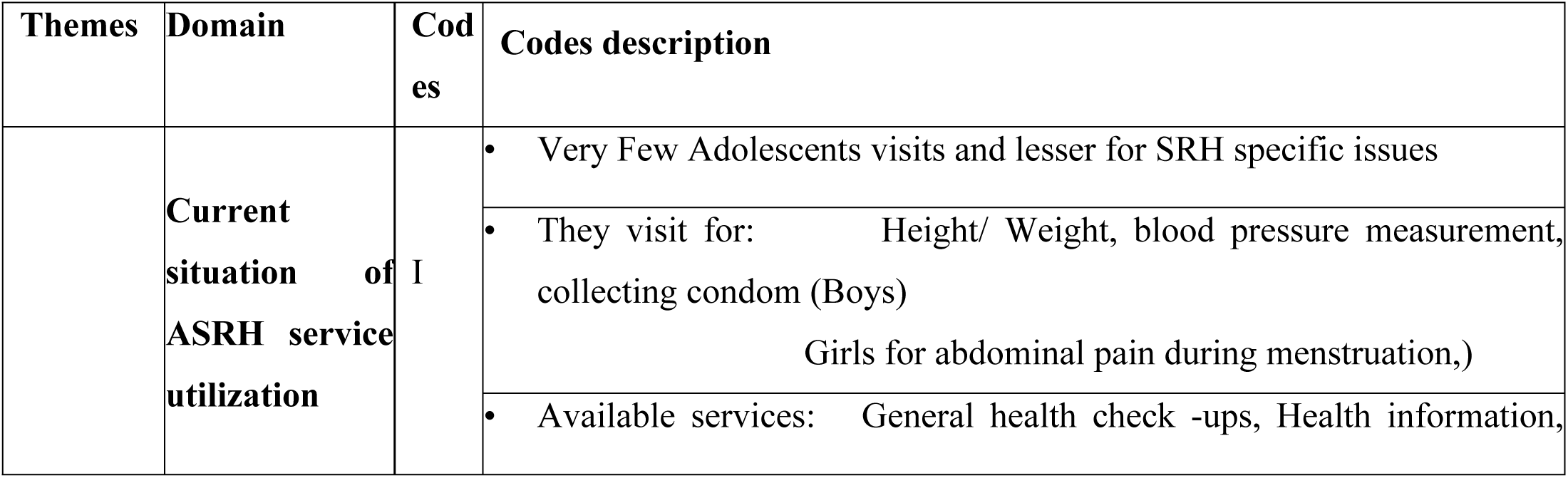
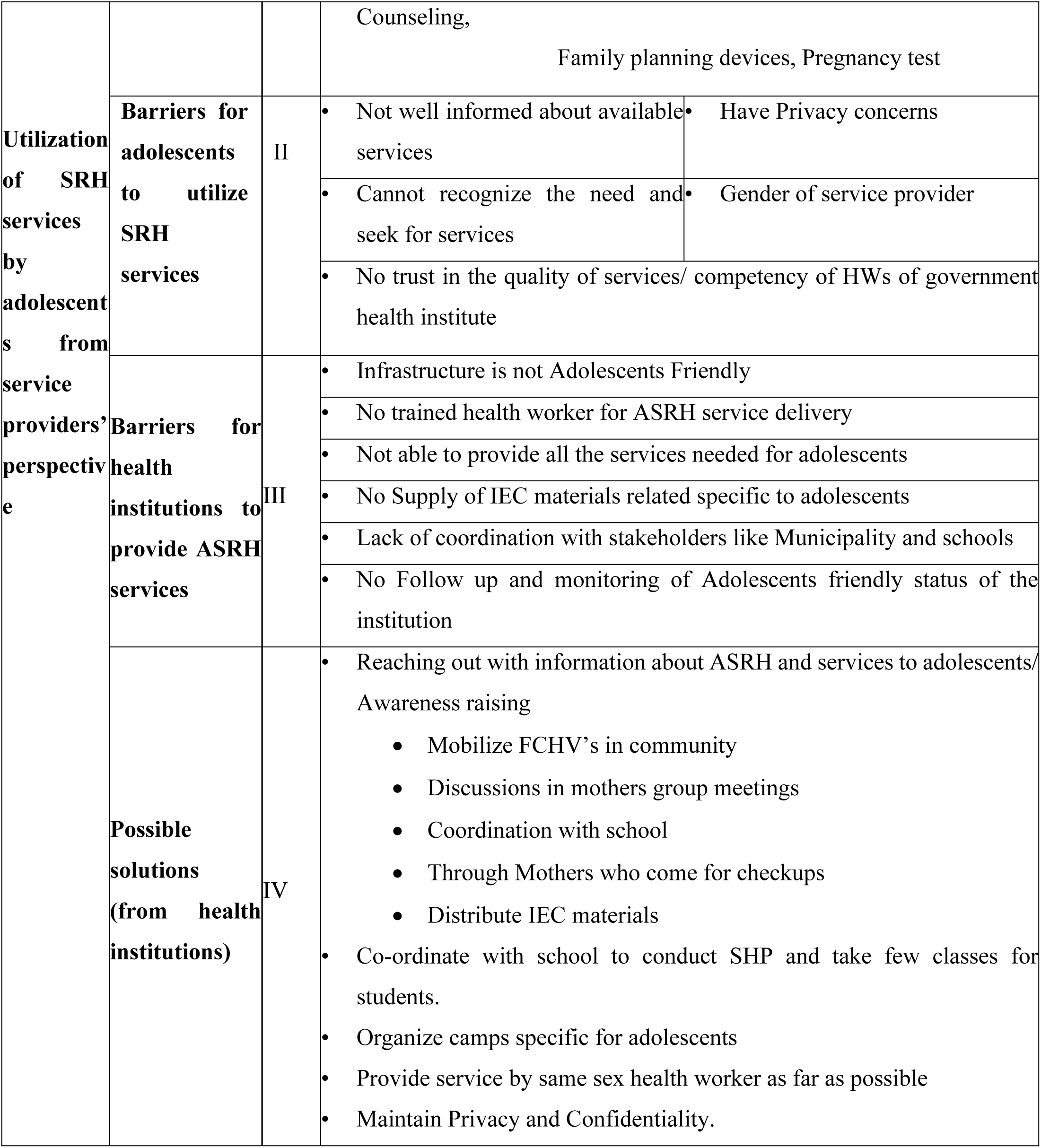
List of codes obtained on Thematic Analysis.

## Discussion

This study shows that respondents who were either married or were in a relationship were 2.202 times more likely to utilize ASRH services than the respondents who were single (Crude OR=2.202, 95% C.I= 1.096, 4.423) more likely to utilize ASRH services than those who were single. It is complemented by studies in the Bhaktapur and also in Surkhet among secondary school students (10-24 years), where 77.08% of the married youths benefitted from the services more than the unmarried youths.(13),(14) In Bhaktapur, it was observed that a married person was six times more likely to utilize ASRH services than unmarried persons (Crude OR: 6.51).(13) This may be due to the fact that there are stigma attached with use of SRH related services by young and unmarried individuals but for those who are married or in relationship, they are more sexually active and their needs for SRH services increase resulting in the use of such services.

This study identified that the respondents who had conversations with their parents about ASRH were 2.308 times more likely to utilize ASRH services than who did not which is comparable to a study in Ethiopia, where those who had a parental discussion on VCT services were ten times more likely to utilize the service compared to those who had no parental communication.(15) However, it contradicts the study among youths in Kathmandu where respondents having distinct interaction with parents were 2 times more likely to utilize SRH services than those having close interaction (OOR: 2.424). (16) The difference may have been due to the difference in the nature of participants in both studies as study in Kathmandu included participants up to 24 years age who may have been able to independently decide to utilize services and did not need communication with parents.

Around three-quarters (77.40%) of them knew about any health institutions that provided ASRH services in their area and more than two third of them had heard about AFHS. The respondents who were aware about these were more likely to use services compared to the others. It concedes with other studies in Bhaktapur and Dang where there was a significant association too.(13),(17) This is because lack of information about the kind of ASRH services available is one of the important barrier to utilizing services, so having any awareness about ASRH or AFHS would be enabling factor towards acquiring those services. For the many respondents (68.94%), the nearest health institution was less than 30 minutes walking distance from their home. Even though distance was not identified as major barrier to utilization of services in this study, other studies in Bhaktapur and Dhading has identified it as an important aspect of accessing ASRH services.(13),(18)

Almost half of the respondents reported that they had felt the need for ASRH services until now, which similar to study in Pyuthan (44%) but is very high compared to the study in Bhaktapur (15%) among participants within the last 12 months.(19),(13)The figure is higher in this research as it collects the information of their needs in their entire life whereas the one in Bhaktapur only takes into consideration the last 12 months from the time of data collection. Two third (66.59%) of the respondents thought they would use the service in the future if needed which is almost double than that of study in Pokhara (34.70%) and Bhaktapur (37.00%).(20),(13)

This study found that the overwhelming majority of adolescents were not utilizing SRH services. Only 13.22% of participants have ever used SRH services to date. This finding is supported by the qualitative study of this research too. In the KII, the health workers mentioned that the service utilization by adolescents is very low, and those specific to SRH-related issues are very rare. Likewise, in the studies of similar nature conducted in Bhaktapur in 2015 and Pokhara in 2018, the utilization was only 9.2% and 17% respectively.(13),(20)Similarly, the household surveys in Kathmandu valley among the youths (15-24 years) and Bhaktapur district among adolescents (10- 19 years) in 2020 also show low utilization of services which is 23% and 24.7%.(21),(22) A recent survey conducted in three states across India found that young men and women (aged 15–24 years) seldom consulted CHWs for issues related to sexual health.(23) The service use is lower in this study than that demonstrated in another study conducted in Pyuthan (44.0%), and also in Myanmar where around two-thirds of youths used some RH services at least once in past.(19),(24) This difference might be due to a different study setting.

In this research, the primary reason for not using services was because adolescents did not think the service was needed (60.39%), which is comparable to the study conducted in Puthyan district, where 56% revealed that they did not feel any sexual and reproductive health related problem; hence they did not go to the health facility.(19) Also, in KII of this study, all health workers mentioned that due to the immature age of adolescents and the multiple changes occurring in their bodies at that age, they are incapable of identifying when they need to get services from health institutions.

Another important reason according to adolescents was that they did not know exactly where to get services. This barrier were acknowledged by services providers too in KII, who mentioned that the adolescents did not visit the health institution because they did not have adequate information about what kinds of services are available at the health institution and adolescents would also doubt that their problems can be addressed in local level government health institutions. Lack of knowledge and information about AFHS was one of the most commonly reported barriers to service utilization by adolescents in the study conducted by UNFPA in Nepal in 2015. (25)

From adolescent’s point of view, other reasons for not utilizing ASRH services were being shy to ask for services, fear of parents, and the opposite gender of the service provider. In KII, service providers also mentioned that at the health institutions, the sex of the service provider would determine how comfortable the adolescents were to share the problems and could also avoid coming to a health facility if the service provider were of the opposite gender. Similarly, around 43.64% of the participants who used the services were unsatisfied with the level of privacy maintained which is also realized by service providers in KII as an important concern because of which, adolescents would rather go to private institutions or institutions that were far from their locality than be seen going to local health institutions by the community people. Research by Upadhyay P on sexual and reproductive health services: utilization pattern of adolescents in Nepal mentioned female adolescents felt embarrassed to express their sexual health problems to male health workers, and the male had similar experiences with female health workers.(19) A journal article published in KMUJ in 2008: Sexual and reproductive health status among young peoples in Nepal: opportunities and barriers for sexual health education and services utilization also reveals similar reasons like fear of parents, unawareness about the benefits of the services, and lack of information.(4) Poor sexual and reproductive health knowledge, lack of privacy, experiences of shame, and healthcare provider attitudes caused barriers to utilizing SRH services by adolescents.(26) Studies in Nigeria and India showed that the majority of adolescents who contracted STI or other reproductive health problems did not use reproductive health services for several reasons, including shame and embarrassment, negative provider attitudes, perceived lack of Confidentiality, fear of being ridiculed, and discrimination they face by some health service staff in accessing contraceptive, sexually transmitted infection services or other services.(27)

The barriers identified in the delivery of services to adolescents were the unavailability of adolescent-friendly infrastructure such as separate rooms or proper waiting spaces, the unavailability of ASRH-trained health workers, and inadequate availability of necessary supplies. A similar finding was found in Kaski where the poor physical infrastructure of health institutions was expressed as one of the barriers by all health workers while providing services and the majority of health workers also expressed their dissatisfaction with not receiving appropriate training.(28) In a qualitative study conducted in Dhading district in Nepal, among adolescents and health care providers, the shortage of ASRH-trained Health Care Providers in health facility which resulted in the allocation of non-ASRH-trained HCPs, were identified as an important factor compromising the quality of health care provided to adolescents.(18) Poor sexual and reproductive health knowledge, the lack of youth-friendly services, lack of confidentiality of services, experiences of shame, and healthcare provider attitudes also have an influence on SRH service utilization by adolescents.(26) A study conducted in Colombia in 2015 to evaluate youth-friendly health services, concluded lack of trained health workers in sexual and reproductive health and the high instability of health workers have a negative impact on the provision of services.(29) Another barrier was the inability of health institutions to provide comprehensive services as they were capacitated to provide only limited services needed. A review article published in 2008, reported that current sexual and reproductive health services in Nepal do not cover a whole range of SRH services that adolescents need and they need to refer to the higher level health facility.(4) A study conducted among 338 higher secondary school adolescents in Bhaktapur reported that 30% of participants feel that currently available services are inadequate to meet their SRH needs.(13)

While observing the health institutions there was no supply of IEC materials specifically designed for ASRH, none of them had displayed the AFS logo, reporting in AFs format was null, and involvement of the adolescent in HFOMC was not in practice. Even though those health institutions were once certified as AF institutions, due to a lack of proper monitoring and follow-up, they were not able to maintain that status anymore. The situation was similar in the observation of four health institutions in Kaski.(28) National Adolescent Sexual and Reproductive Health Programme: Mid-Term Evaluation Report, 2013, mentioned that ‘AFS logo signboard’ is helpful to attract adolescents as it informs adolescents that services are available.(30)

## Conclusion

Most of the participants were 15-17 years old, female, and were studying in class 12. Very few were married and few were in a relationship. About one-third of the participants had a conversation with the parents regarding different matters of ASRH. Many knew about any health institutions that provided ASRH services in their area. The nearest health institution was less than 30 minutes walking distance from the home for the majority of respondents. Only a few had heard about AFHS. Even though many had felt the need for ASRH services in their lifetime, many were still unsure about using services in the future; few were sure they would not use them. The utilization of ASRH services during their lifetime was very low. The main reason was that the adolescents thought it was not required to go to health institutions for their problems.

Overall, poor utilization of SRH services by adolescents was due to the inability of adolescents to identify their health needs as a result, thinking that they do not need health services, being uninterested in using such services, being ashamed to ask for services, did not know exactly where to get them, poor availability of AF health institutions and services, inability to provide comprehensive services, inadequate availability of necessary supplies, lack of trained human resources and so on. So, to increase the utilization, awareness should be generated among adolescents of their ASRH needs and available ASRH services in health institutions through mutual coordination with schools because it is their vital source of information. Similarly, adolescent-friendly infrastructure and environment and trained health personnel should be managed to provide services.

## Data Availability

All the data generated in the study are included in the manuscript.

## Acknowledgements

Immense Gratitude towards the Health coordinator of Godawari Municipality, Mr. Rewati Raj Karki for his support to conduct the research among health institutions of the Municipality as well as all the schools for their support. The support from all the members of faculty of public health, Manmohan Memorial Institute of Health Sciences is also appreciative. Above all, cordial thanks to all the respondents for their time without whom this study would not have been possible.

